# Carnosine supplementation improves glucose control in adults with pre-diabetes and type 2 diabetes: a randomised controlled trial

**DOI:** 10.1101/2023.03.18.23287432

**Authors:** Rohit Hariharan, James Cameron, Kirthi Menon, Jakub Mesinovic, Paul Jansons, David Scott, Zhong X Lu, Maximilian de Courten, Jack Feehan, Barbora de Courten

## Abstract

Type 2 diabetes (T2DM) is a major cause of morbidity and mortality globally. Carnosine, a naturally occurring dipeptide, has anti-inflammatory, antioxidant, and anti-glycating effects, with preliminary evidence suggesting it may improve important chronic disease risk factors in adults with cardiometabolic conditions. In this randomized controlled trial, 43 adults (30%F) living with prediabetes or T2DM consumed carnosine (2 grams) or a matching placebo daily for 14 weeks to evaluate its effect on glucose metabolism assessed via 2 hr, 75g oral glucose tolerance test. Secondary outcomes included body composition analysis by dual energy x-ray absorptiometry (DEXA), calf muscle density by pQCT and anthropometry. Carnosine supplementation decreased blood glucose at 90 minutes (−1.31mmol/L; p=0.02) and 120 minutes (−1.60mmol/L, p=0.02) and total glucose area under the curve (−3.30mmol/L; p=0.04) following an oral glucose tolerance test. There were no additional changes in secondary outcomes. The carnosine group results remained significant before and after adjustment for age, sex, and change in weight (all>0.05), and in further sensitivity analyses accounting for missing data. There were no significant changes in insulin levels. Likely mechanisms may include changes to hepatic glucose output explaining the observed reduction in blood glucose without changes in insulin secretion following carnosine supplementation. This study provides preliminary support for larger trials evaluating carnosine as a potential treatment for prediabetes and the early stages of T2DM.

## Introduction

Type 2 diabetes (T2DM) continues to place an immense burden on healthcare systems globally^1^. The World Health Organization (WHO) estimates that over the last 40 years, the number of people with diabetes has increased from 108 million to 451 million and will increase to 693 million by 2045^1,2^. This rapid increase has cemented diabetes as a leading cause of mortality^3^ and a major contributor to burden of disease globally^4^ and a leading cause of mortality^3^. Diabetes increases the risk of all-cause mortality by 2-3 times^5^ and contributes to as much as 80% of all premature deaths caused by non-communicable disease^5^. It is also an immense economic burden, with $760 billion USD spent globally on diabetes and its complications in 2019 alone^5^. This is projected to rise to $825 billion by 2030 and $845 billion by 2045^5^.

Due to its significant healthcare and societal burden, effective management of T2DM is critical. The principal aims of management of T2DM are to lower and stabilize blood glucose levels, which decreases the incidence and severity of microvascular and macrovascular complications, improves quality of life, and reduces mortality^6^. More than 120 million people with diabetes worldwide are prescribed metformin as their main therapy, making it the most broadly used glucose-lowering medication globally^7^. Other medications used include dipeptidyl peptidase 4 (DPP4) inhibitors, sodium-glucose co-transporter 2 (SGLT2) inhibitors, sulfonylureas, and the glucagon like peptide (GLP-1) receptor agonists^8^. These medications are hampered by adverse events, including hypoglycemic episodes, with resultant sequelae, and some cannot be prescribed in patients with common diabetic comorbidities, such as severe chronic renal disease^9^. This makes the identification of safe and effective means to better control glucose an ongoing priority for researchers.

Carnosine, anserine, and ophidine/balenine, a group of naturally occurring histidine containing dipeptides (HCD), are present in large amounts in the skeletal muscle and brain of mammals^10-14^. Carnosine has several physiological activities in humans, being a potent antioxidant, and metal chelator as well as anti-inflammatory, and anti-glycating effects^14,15^. Carnosine supplementation has been shown to improve glycemic control and insulin levels in adults with prediabetes and diabetes^16-19^, and it also has a positive effect on the blood lipidome by maintaining levels of trihexosylceramide and phosphatidylcholine and free cholesterol levels^17,20-22^.

However, studies investigating the role of carnosine in glucose metabolism are limited by small numbers of participants, frequently treated concomitantly with a variety of glucose-lowering medications as well as a lack of randomized placebo-controlled studies of participants with overweight or obese. Therefore, our aim was to evaluate the effect of a 14-week carnosine supplement regimen for T2DM and make direct measurements of glucose metabolism in adults with prediabetes or T2DM. We hypothesize that oral carnosine supplementation for 14 weeks would improve glucose metabolism and reduce insulin levels in patients with prediabetes or T2DM compared to placebo. A recent systematic review and meta-analysis of 20 studies determined that b-alanine, a precursor to carnosine, had no effect on body composition in athletes ^23^, however it is unknown whether it has any effect in patients with cardiometabolic disease. In our trial, we aim to utilize DXA to further elucidate the effect of carnosine supplementation on whole body mass, fat percentage, fat mass and fat free mass in group of individuals with overweight but not obese.

## Methods

### Study design

The study was a parallel-group double-blinded randomized control trial (RCT). Informed written consent was provided by all participants before commencing the trial. The trial was registered at clinicaltrials.gov (NCT02917928), and was conducted according to the Standardized Protocol Interventions: Recommendations for Interventional trials^24^ and followed the CONSORT guidelines^25^. The study was approved by Monash Health ethics committee (Ref: 16061A). and conforms to the Declaration of Helsinki^26^.

### Sample size

The sample size of the study was calculated using the G-Power software, with blood glucose concentrations via oral glucose tolerance test as the primary outcome. Data from a sub-study in this trial on subjects with overweight or obesity^27^, and our trial in a cohort of participants with overweight or obesity^16^, and from a study of subjects with diabetes by our group(ref); who had mean HbA1c of 7 and fasting glucose of 10 was considered in the calculation. A sample size of 22 in each arm was required to detect a 20% change in fasting glucose and absolute change of 0.5 in HBA1c for 80% power, with target recruitment set to 50 participants based on a type I error of 0.05 (two-tailed)^28^. A 20% change in insulin sensitivity was selected based on evidence of similar improvement in insulin sensitivity when treated with troglitazone, an hypoglycemic medication and insulin sensitiser^27^.

### Study population

To be eligible, participants were required to be aged 18-70 years. All participants had pre-diabetes or T2DM diagnosed by OGTT (according to the WHO guidelines) and were untreated, or taking only metformin. Participants were excluded if their HbA1c was 8% or higher or had changed their medications in the past 3 months. Participants had to have BMI less than 40 kg/m^2^ and a stable body weight (no more than 5kg above or below the current weight over the last 12 months). Participants were otherwise without diagnosis of any renal, cardiovascular, hematological, respiratory, gastrointestinal, or central nervous system disease, and not pregnant or lactating. Participants were not required to change their diets or routine physical activity but were not included if they consumed more than 4 standard drinks per week for men and 2 standard drinks per week for women or if they smoked. Recruited participants were required to remain on the same medications during the 14-week trial. The participant would be discontinued from the study if their HbA1c became greater than 8% and a medication change was stipulated by their medical practitioner at any stage of the study.

### Recruitment, Intervention and Randomization

Participants were recruited through advertising via poster, radio, email newsletters and newspapers. Interested participants were first contacted via phone and completed a screening eligibility questionnaire. Potentially eligible participants were then invited for a screening visit, where they were assessed against the inclusion and exclusion criteria, underwent anthropometry and a screening OGTT test at the Clinical Trials Centre of the Monash Health Translational Research Precinct. Eligible participants were randomized to intervention or placebo; the randomization used block sizes of 4 by sex, using computer-generated randomization codes sent to the clinical trials pharmacy by the study statistician. The carnosine group were asked to take 2g of carnosine (Flamma Group, 1g twice daily including the study days) and the placebo group received an equivalent amount of methylcellulose (1g twice daily) for 14 weeks. Medication was dispensed in indistinguishable capsules and containers to ensure participants and researchers were blinded to the group allocation. Participants were asked to inform the trial doctor of any adverse events as they arose and the trial doctor followed up with all participants at the end of the trial to discuss these. Participants were asked to return the treatment containers at the end of the study to assess treatment compliance. Compliance was evaluated by counting number of unconsumed capsules and dividing by number of capsules provided.

### Outcomes

The primary outcome was plasma glucose level on 2-hour oral glucose tolerance test (OGTT). Secondary measures included HbA1c, insulin levels, body composition, calf muscle density, and anthropometric parameters. All outcomes were measured at baseline and repeated after 14-weeks of intervention.

### Anthropometric, biochemical and body composition assessment

The study physician collected participants’ medical history in person and performed a physical examination. This included collecting anthropometric data using a digital scale (Tanita BWB-600) and standing stadiometer, and vital signs collected via Omron digital blood pressure monitor (Model: BBP-742). Waist and hip measurements were measured using a measuring tape at the midpoint between the upper iliac crest and the lowest rib, and at the widest part of the buttocks. Participants’ bloods were collected after an overnight fast of at least 10 hours using aseptic methods, and analyzed by the National Association of Testing Authorities accredited Monash Health pathology service, which operates an automated core laboratory at Monash Medical Centre Clayton.

Whole-body dual-energy x-ray absorptiometry (DXA; Hologic Discovery A, Hologic, USA) was used to estimate fat and lean mass. The DXA scanner was calibrated daily with the manufacturer’s spine phantom and the short-term intra-individual coefficient of variation (CV) for lean and fat mass was 1.60% and 2.67%, respectively. All DXA scans were analyzed by the same investigator.

A single 2.5 mm transverse peripheral quantitative computed tomography (pQCT) scan (Stratec XCT3000, Stratec Medizintechnik GmbH, Pforzheim, Germany) at a speed of 20 mm/s and voxel size of 0.8 mm was obtained at 66% of the tibial length of the non-dominant leg. The length of the tibia was determined by measuring the distance between the prominence of the medial malleolus and the tibial plateau. A planar scout view of the distal tibia was used to locate scan sites, and reference lines were placed parallel to the distal joint surface of the tibia. Calf muscle density (mg/cm^3^; density of tissue within the muscle compartment after removal of subcutaneous fat and bone) was estimated using manufacturer’s algorithms and software (version 6.2). Scans were analyzed using a smoothing filter (F03F05) at a threshold of 41 mg/cm^3^. The device was calibrated daily using the manufacturer’s phantom and the short-term intra-individual CV for muscle density was 1.2%. All pQCT scans were analyzed by one individual. Adverse events while participating in the 14-week intervention period were recorded in the progress notes as they arose and were reviewed with the chief investigator at the start of every year while preparing the trial progress reports.

### Glucose and insulin assessment

Participants completed a 2-hour OGTT following overnight fasting. The OGTT drink containing 75g of glucose was consumed within 5 minutes after which bloods were collected aseptically into tubes (Serum Separation Tubes – SST II) at 30-minute intervals (0, 30, 60, 90, 120 mins). After centrifugation, the serum samples were stored at -80°C until analysis. Serum samples at all the time points were measured for glucose and insulin, with the values also used to calculate the area under the curve (AUC) by the trapezoidal method. The homeostatic model assessment of HOMA-IR (insulin resistance), HOMA-β (steady state β-cell function %) and HOMA-S% (Insulin sensitivity) were also calculated as described previously^16,28,29^.

### Statistical Analysis

Statistical analysis was performed with the statistical softwares JASP (v0.16.3), and SPSS (v29.0.0.0). Parametric data were reported as mean and standard deviations and non-parametric data were reported as median and interquartile range. Skew was assessed by Shapiro-Wilk testing, with skewed data logarithmically transformed with satisfactory transformation assessed by evaluation of residual plots. Continuous variables were analyzed using Analysis of Covariance (ANCOVA) adjusted for baseline values. Additional ANCOVA was performed with diabetes (prediabetes vs T2DM) status, obesity status (BMI > 30) or metformin intake as covariates to evaluate interaction effects. All tests were two-tailed, and alpha level of significance was set at 0.05. The primary analysis was performed on a complete case protocol; however, an additional sensitivity analysis was performed with multiple imputation on missing cases was performed. Missing data was imputed with predictive mean matching, with a total of five imputations modelled. The resulting data sets were analyzed as above, with the pooled mean and standard error of the imputations, and the range of the resulting p-values reported.

## Results

A total of 88 participants were assessed for eligibility, with 49 eligible for enrolment and randomization. A total of 43 participants (13 women years and 30 men completed the trial at 14 weeks (23 placebo and 20 carnosine). Of 43, 22 participants were assessed as pre-diabetic and 21 as diabetic based on their screening OGTT. Five participants were lost to follow-up (Two placebo, three carnosine), and one additional participant was withdrawn after initiating treatment with an additional medication following randomization (Figure 1). Baseline participant data is presented in table 1.

**Figure 1:**
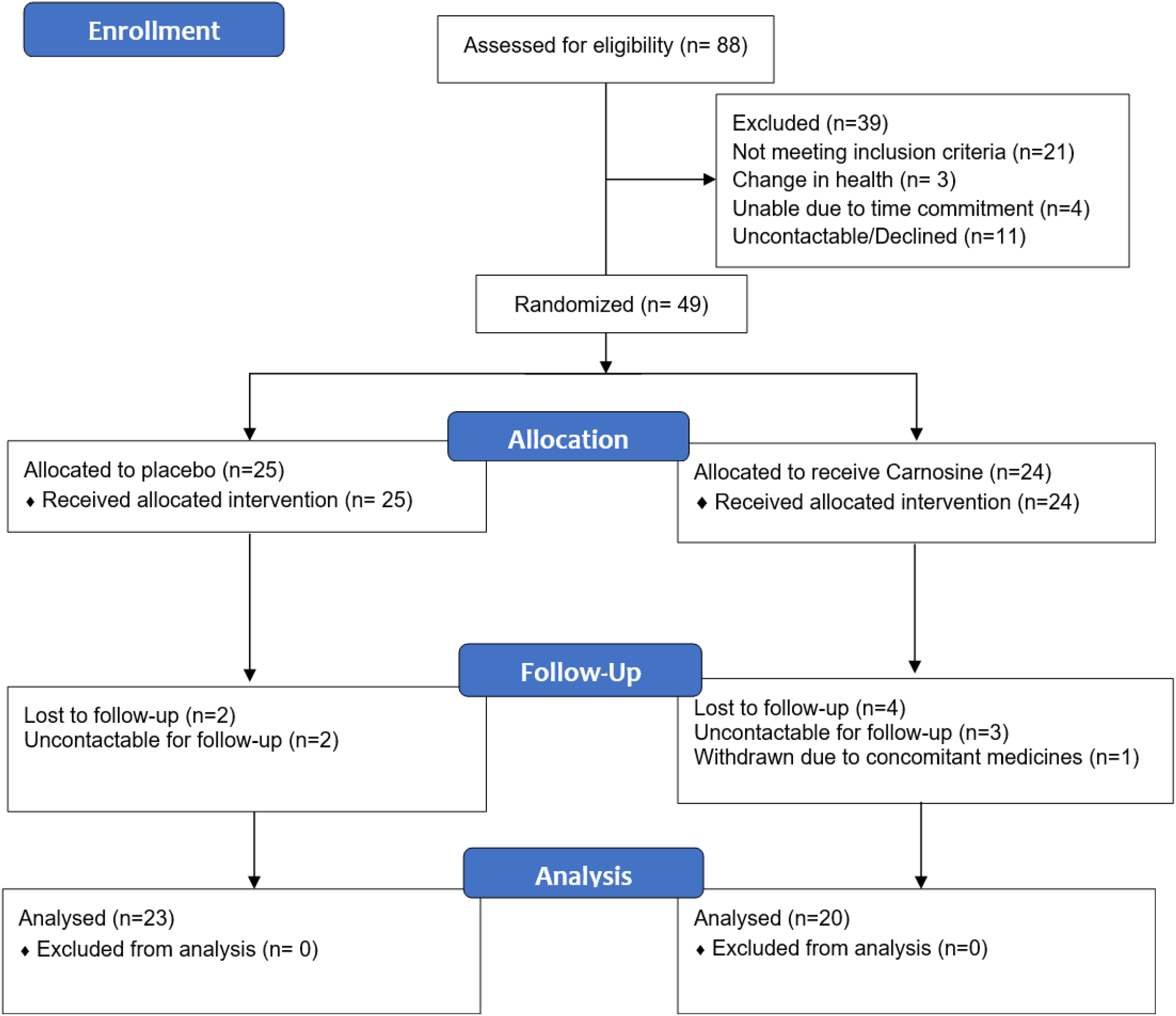
Study flow diagram.

**Table 1:** Baseline characteristics of participants

|  | Placebo (n=23) |  | Carnosine (n=20) |  |
| --- | --- | --- | --- | --- |
| Age – years,<br>(median [IQR]) | 52 [28.4 – 65.3] |  | 53.8 [30.1 – 66.3] |  |
| Female, (n [%]) | 7 [30%] |  | 6 [30%] |  |
| Pre-diabetic/Diabetic | 11/12 |  | 11/9 |  |
| Treated with Metformin | 9 |  | 8 |  |
|  | Mean | SD | Mean | SD |
| Weight (kg) | 81.8 | 14.5 | 88.4 | 23.8 |
| Height (cm) | 169.0 | 10.3 | 172.3 | 12.3 |
| Waist Circumference (cm) | 99.0 | 8.8 | 103.3 | 14.7 |
| Hip Circumference (cm) | 102.5 | 7.9 | 105.7 | 15.1 |
| Glucose - 0mins (mmol/L) | 6.4 | 1.1 | 6.4 | 1.1 |
| Glucose - 30mins (mmol/L) | 9.6 | 1.3 | 10.3 | 2.3 |
| Glucose - 60mins (mmol/L) | 12.1 | 2.1 | 13.0 | 2.4 |
| Glucose - 90mins (mmol/L) | 12.3 | 2.7 | 13.3 | 3.5 |
| Glucose - 120mins (mmol/L) | 11.9 | 3.0 | 12.0 | 3.5 |
| Glucose AUC | 43.2 | 7.5 | 45.8 | 9.792 |
| Insulin - 0 mins (mIU/ml) | 9.7 | 5.7 | 14.6 | 19.3 |
| Insulin - 30 mins (mIU/ml) | 32.7 | 30.3 | 34.4 | 17.3 |
| Insulin - 60 mins (mIU/ml) | 41.8 | 26.9 | 67.5 | 58.0 |
| Insulin - 90 mins (mIU/ml) | 56.1 | 42.3 | 96.5 | 92.0 |
| Insulin - 120 mins (mIU/ml) | 70.9 | 64.5 | 104.0 | 111.8 |
| Insulin AUC | 159.1 | 113.3 | 257.5 | 216.6 |
| HbA1c (%) | 6.7 | 0.8 | 6.5 | 0.6 |
| HOMA-IR | 2.7 | 1.7 | 4.1 | 5.3 |
| HOMA-S | 51.7 | 37.6 | 61.4 | 101.8 |
| **HOMA- $\beta$** | 80.8 | 63.6 | 118.8 | 173.3 |

### Effect of carnosine supplementation on obesity measures

After the 14-week intervention, we detected a decrease in BMI of 0.74 kg/m^2^ (p=0.048) in the carnosine group compared to placebo; but this did not remain significant when adjusting for diabetes status, obesity status or by metformin intake, and did not show any interaction effects with these variables. There were no differences in body weight, waist circumference or waist-to-hip ratio, body fat percentage (BF%), fat mass and fat free mass between the groups (all p>0.05) (Table 2. There were no interaction effects with diabetes status, obesity status or by metformin intake.

**Table 2:**
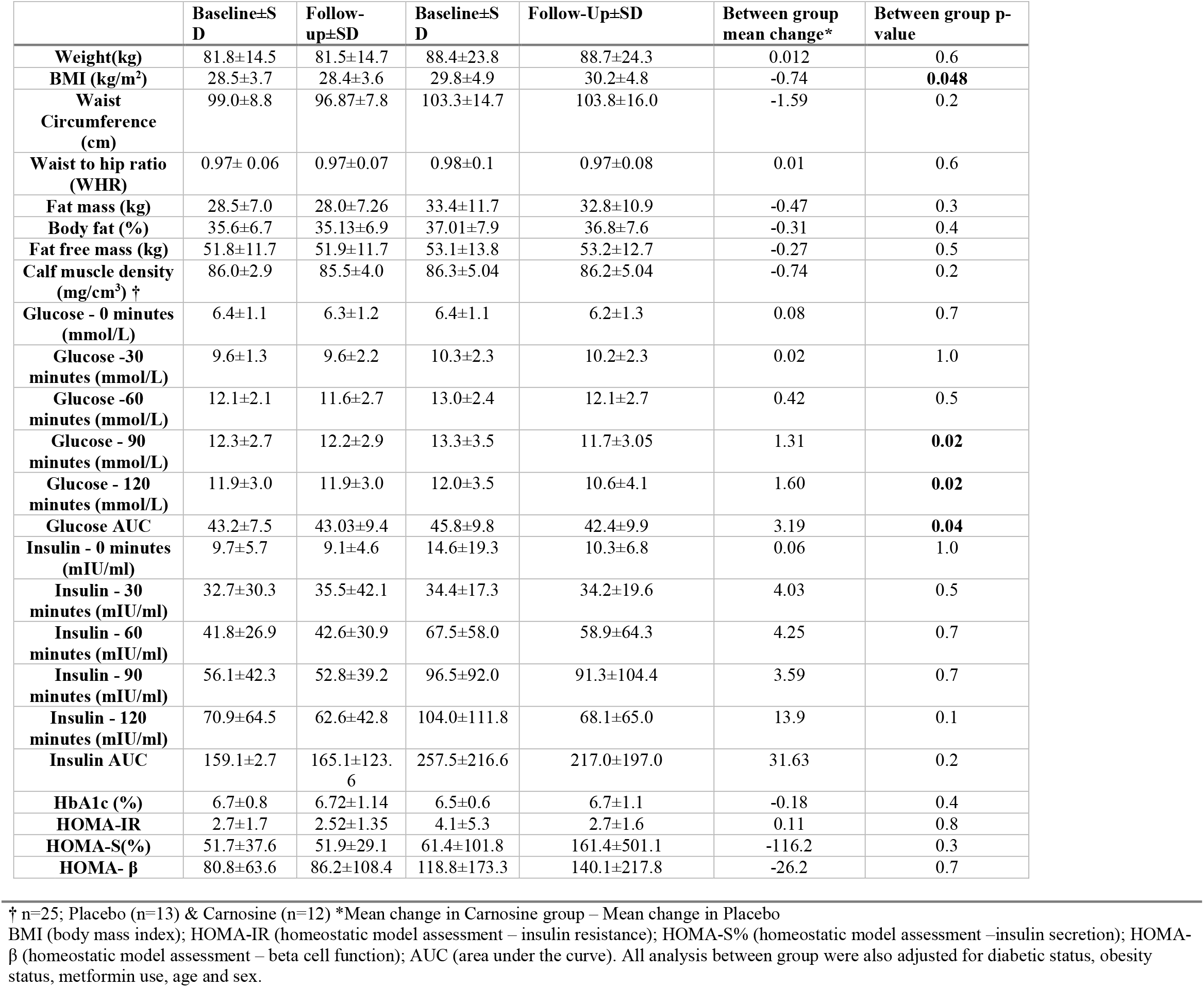
Baseline and follow-up values and changes in anthropometric, body composition and metabolic measures after 14 weeks of supplementation

### Effect of carnosine supplementation on glycemic measures

Carnosine supplementation reduced glucose levels at 90-minutes (−1.60 mmol/L, p=0.018) and 120-minutes (−1.40 mmol/L, p=0.016), as well as glucose AUC (−3.30 mmol/L, p= 0.04) compared to placebo controls (Table 2, Figure 2). There were no changes in any other glucose or insulin measures, insulin AUC, HOMA-S, HOMA-β or HOMA-IR (all p>0.05) (table 2). There was no interaction between changes in any of the variables and diabetic status, obesity status or by metformin intake. The glucose results remained consistent in sensitivity analysis evaluating the effect of missing data imputation (Table 3).

**Table 3:** Sensitivity analysis of glucose and insulin results, through multiple imputation of missing results, and resulting ANCOVA. Five imputations were used with pooled means and standard errors, and the resulting p-value ranges provided.

| Variable | Pooled imputed mean estimates (Std Error) |  | Range of imputed p-values |
| --- | --- | --- | --- |
| **Glucose - 0 minutes (mmol/L)** | *Placebo* | 6.23 (0.96) | 0.19-0.97 |
|  | *Carnosine* | 6.21 (0.71) |  |
| **Glucose -30 minutes (mmol/L)** | *Placebo* | 10.25 (0.63) | 0.25-0.99 |
|  | *Carnosine* | 9.86 (0.55) |  |
| **Glucose -60 minutes (mmol/L)** | *Placebo* | 11.90 (0.80) | 0.18-0.88 |
|  | *Carnosine* | 11.66 (0.55) |  |
| **Glucose - 90 minutes (mmol/L)** | *Placebo* | 12.69 (0.46) | 0.010-0.018 |
|  | *Carnosine* | 11.31 (0.41) |  |
| **Glucose - 120 minutes (mmol/L)** | *Placebo* | 12.05 (0.53) | 0.010-0.023 |
|  | *Carnosine* | 10.56 (0.48) |  |
| **Glucose AUC** | *Placebo* | 44.06 (1.03) | 0.024-0.055 |
|  | *Carnosine* | 41.12 (1.05) |  |
| **Insulin - 0 minutes (mIU/ml)** | *Placebo* | 9.68 (1.12) | 0.68-0.96 |
|  | *Carnosine* | 9.80 (1.20) |  |
| **Insulin - 30 minutes (mIU/ml)** | *Placebo* | 33.52 (9.68) | 0.08-0.76 |
|  | *Carnosine* | 21.78 (13.68) |  |
| **Insulin - 60 minutes (mIU/ml)** | *Placebo* | 49.03 (9.65) | 0.34-0.86 |
|  | *Carnosine* | 45.69 (9.33) |  |
| **Insulin - 90 minutes (mIU/ml)** | *Placebo* | 63.82 (10.24) | 0.18-0.81 |
|  | *Carnosine* | 77.44 (11.55) |  |
| **Insulin - 120 minutes (mIU/ml)** | *Placebo* | 69.08 (7.94) | 0.086-0.82 |
|  | *Carnosine* | 63.15 (11.17) |  |
| **Insulin AUC** | *Placebo* | 185.32 (19.84) | 0.42-0.99 |
|  | *Carnosine* | 181.87 (21.61) |  |

**Figure 2:**
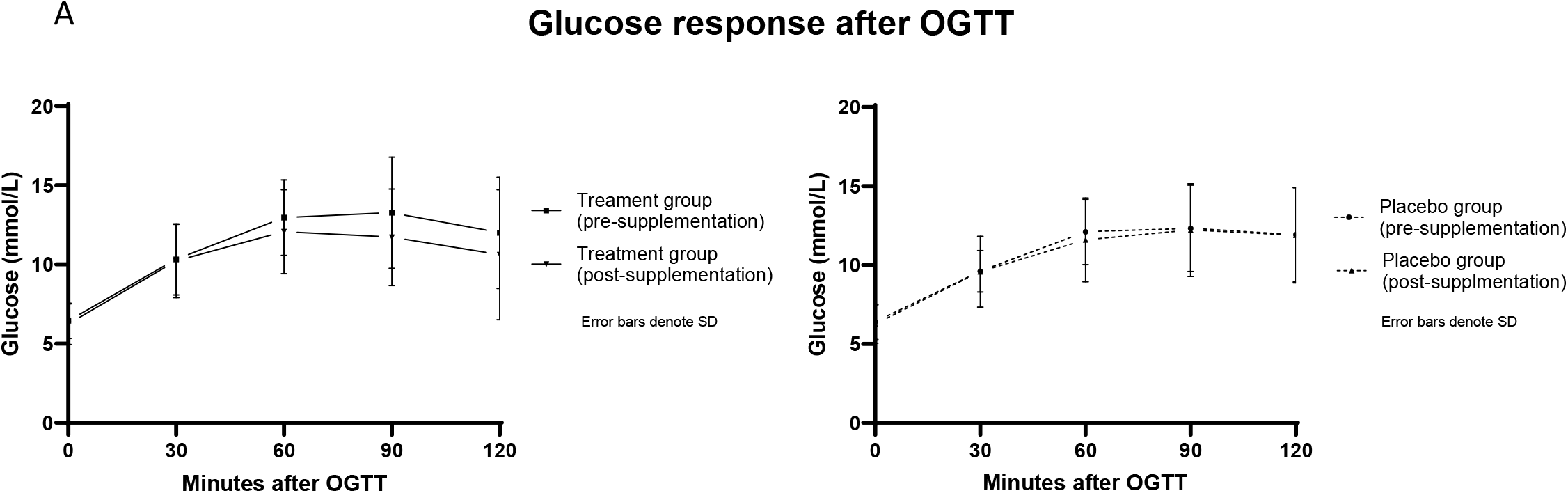
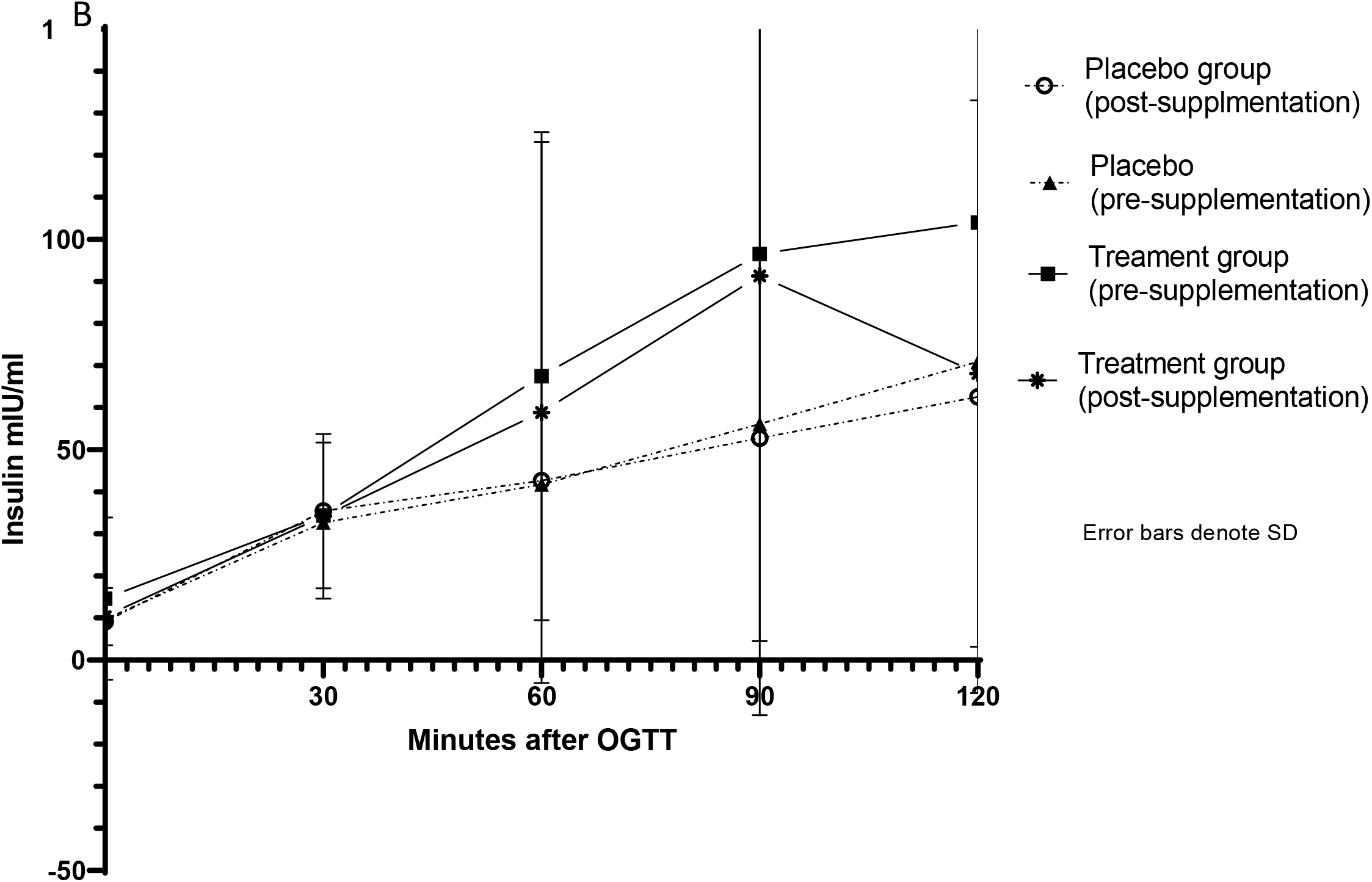
Glucose levels (A) and Insulin level (B) after oral glucose tolerance test at baseline and after 14-week intervention by intervention group.

**Figure 3:**
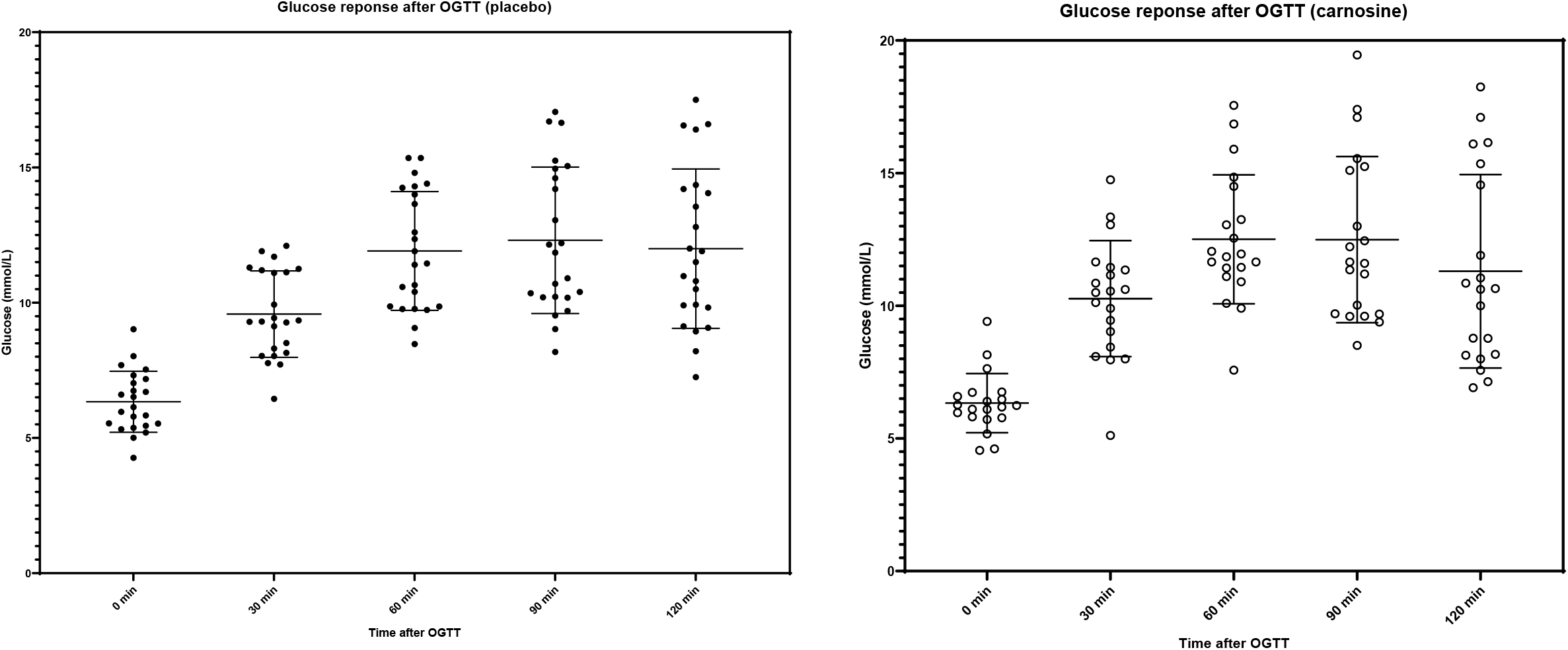
Glucose response after OGTT per participant. Bars denote Mean and SD.

Carnosine supplementation did not result in any adverse events, with the only reported adverse event being pain and mild bruising at the site of venipuncture in one participant. Overall, compliance was excellent (100%) with all participants consuming all capsules given to them.

## Discussion

In our randomized controlled study, we showed that carnosine supplementation for 14 weeks reduced 90 and 120-minute glucose levels after OGTT, and reduced glucose AUC. There were otherwise no significant changes to body composition, nor other glucometabolic measures. Given carnosine is safe, cost effective^30^, and available over the counter in most countries, the reductions in glucose levels make carnosine an attractive agent for the management of prediabetes and diabetes, and warrants further large-scale trials to evaluate its long-term effects in the prevention and treatment of T2DM.

In this trial, the reduction of 3.3 mmol/L in glucose AUC following a OGTT represents a 7.2% decrease in glucose response, which is likely to provide clinically significant outcomes in both preventing transition from prediabetes and in diabetic complications, particularly given 40% of the cohort was already using metformin^31^. In the Diabetes Prevention Program Outcomes Study (DPPOS), participants with prediabetes who returned to normoglycemia had 56% lower risk of diabetes than those who remained prediabetic^32^. Similarly, our recent meta-analysis pooling 23 interventional studies showed improvement in glycemic control for participants with both type 1 and T2DM, following carnosine supplementation leading to improvement of fasting glucose (mean difference (MD) [95% CI]= -0.6 mmol/L [-1.1, -0.1], p=0.03 and HbA1c (MD [95% CI]= -0.5% [-0.4,-0.6], p<0.001) ^33^. In the meta-analysis, sensitivity analyses eliminating studies in which fasting glucose was elevated at baseline, still favored supplementation with carnosine^33^, aligning with the results from the present study. We and others also highlighted that there was insufficient data to evaluate whether carnosine supplementation improved glucose following OGTT^33,34^, although some showed glucose lowering effects in people with diabetes ^17^. While our findings of improved glucose control after OGTT align with these findings, our study did not show changes in HbA1c levels or fasting glucose ^35,36^. This is potentially due to differences in participants, with our study investigating people with pre-diabetes and relatively well-controlled T2DM. Additionally, the 14-week intervention in this study may have been too short to cause an appreciable change in HbA1c, which typically requires longer to decrease. While the evidence provided in this study has demonstrated an effect of carnosine supplementation on glucose concentrations, the mechanisms underpinning this are unclear. The glucose-lowering effect was in the absence of an alteration in insulin concentrations, as well as indirect measures of insulin sensitivity, which further highlighted the differences in baseline insulin levels between the placebo and treatment groups (2.7 mIU/ml vs 4.1mIU/ml); is a possible reason as to why a significant change in insulin at 14 weeks was not detected. This may indicate decreased hepatic glucose output a potential mediator of the effect seen in this trial.

Carnosine is rapidly hydrolyzed by serum carnosinase to its constituents b-alanine and histidine. Carnosine’s poor bioavilaibilty in humans is highlighted by the fact that individuals with CNDP1 genotype characteristed by low serum carnosinase activity are often afflicted by carnosinemia^37^, however carnosinemia can be induced by intake^37^. The high variation in serum carnosinase activity seems a natural variation and depends upon the polymorphism of the CNDP1 genotype inherited.

Although our study found a decrease of 0.74 kg/m^2^ in BMI in the carnosine group (p=0.045) after 14-week supplementation compared to the placebo; with no significant change in weight between follow-up and baseline, this is unlikely to be clinically relevant, and is potentially random. Our study found no change in waist circumference or waist-to-hip ratio after carnosine supplementation, in line with previous studies of similar cohorts^16,23^. Liu et al^36^ reported in one trial that carnosine as part of a combined supplement with cinnamon and chromium increased lean mass, which is in contrast to our trial. Although, our study cohort was largely overweight rather than obese and several participants had been pretreated with metformin and/or diet control and exercise, which could explain the lack of effect observed in these measures. Another meta-analysis^33^, also found a significant 3.5cm decrease in waist circumference, findings not seen here. A prior RCT^38^ compared β-alanine (BA) supplementation only (n=8) (the rate-limiting precursor to carnosine and a preferred method to increase muscle carnosine levels by circumventing serum carnosinase activity), creatine only (n=8) and BA plus creatine (n=7) or placebo (n=7) for 28 days along with a structured exercise routine in a group of physically active females^38^. They found that despite addition of exercise, there were no reductions in body weight, fat mass, body fat and fat-free mass^38^ in any cohort; findings echoed in another trial of β-alanine and exercise combined^39^. Another recent meta-analysis of 426 participants found no effect of supplementation on any measure of body composition, and no interaction effects resulting from resistance training, endurance training or combined training ^23^. This data, and ours provides evidence that neither carnosine nor β-alanine supplementation seems to affect body composition directly, with or without complimentary exercise in adults without obesity. However, it should be noted that most of these studies were in athletes who are less likely to show large changes in body composition. Further trials which supplement carnosine in individuals with obesity may better elucidate an effect of carnosine on body composition.

The findings of the current study agree with previous work demonstrating that carnosine has glucose-lowering effects in individuals with impaired glucose tolerance, however, our data was unable to demonstrate any changes in insulin levels as found in other similar trials^16,40^, possibly due to the large number of participants with diabetes being treated with metformin in our cohorts or the fact that we used indirect measurements for insulin sensitivity and secretion. We have shown previously that carnosine supplementation leads to increases in insulin sensitivity and secretion in individuals with overweight and obesity^16^. Future studies will need to employ gold-standard euglycemic-hyperinsulinemic clamp and intravenous glucose tolerance test to accurately evaluate carnosine’s effect on these measures.

Chronic inflammation, oxidative stress and advanced glycation end products (AGE) and advanced lipid peroxidation end products (ALE) have all been implicated in the pathogenesis of diabetes and its complications ^41,42^. Carnosine is thought to improve glycemic control at least in part via anti-AGE action by preventing the formation of cross-linked proteins by methylglyoxal (MG) and neutralizing it^43,44^. Free sugars undergo a series of biochemical changes^45^ resulting in their oxidization after which they covalently bond to proteins^25^. These modified proteins form permanent cross-linkages with oxidized sugars and other proteins over time to form AGEs, which through activation of immune processes, contribute to worsening hyperglycaemia^46,47^. Carnosine has been proposed to improve a range of cardiovascular, dyslipidemic, and inflammatory outcomes ^15,17,48^ – all essential for the care of patients with diabetes. This adds additional benefits to the use of carnosine as a part of treatment of T2DM, particularly in the light of the glucose lowering effect demonstrated here. The analysis of inflammatory and lipid profiles collected as part of this trial is currently being analysed and will be reported in another article.

### Strengths, Limitations, and future direction

While this study was methodologically robust, there are some limitations We used surrogate measures of insulin sensitivity and secretion HOMA-IR HOMA-S and HOMA-β which are not as sensitive as the gold-standard methods. Our analysis showed that the baseline insulin level for the carnosine group was 29.8% higher than the baseline insulin for the placebo group. This random characteristic as well as missing OGTT and insulin data for a few participants (table 2) may have further obscured the detection of a significant change in insulin levels after 14 weeks, even though the carnosine group lowered HOMA-IR scores to greater degree than the placebo group (change of -1.4 mIU/ml vs -0.18 mIU/ml), although these results were not stastically significant. The higher insulin level in the carnosine group at baseline may have been a reason why a larger therapeutic change was evident after treatment. Moreover, the trial was limited by a smaller sample size due to the difficulty in recruiting participants with prediabetes and T2DM treated with metformin only and furthermore by COVID-19 pandemic which halted the trial for several months. Another limitation was the relatively short follow-up period, which left questions surrounding treatment durability unanswered.

## Conclusion

A 14-week regimen of carnosine supplementation reduced glucose concentrations following OGTT in patients with prediabetes and diabetes but had no effect on insulin concentrations or obesity measures. While this makes carnosine a promising candidate in the management of T2DM, further randomized controlled trials of carnosine supplementation for cardiometabolic conditions are warranted, particularly in the light of carnosine’s additional protective benefits to cardiovascular and inflammatory outcomes relevant to diabetes^33,49^.

## Data Availability

All data produced in the present study are available upon reasonable request to the authors

## Acknowledgements

We acknowledge Flamma Group for providing carnosine for the trial and Ms. Josphine Johnson, Dr. Estifanos Baye, Ms Paula Fudge-Larsen and Dr. Martin Schonn for their assistance with data collection. We also acknowledge funding agencies including CASS and Royal Australasian College of Physicians foundations. Also, we thank volunteers Dr. Aylin Hilberath, Dr. Dilek Tuncel, Dr. Lachlan B McMillan and Dr. Mavil May Cervo for their assistance.

## References

1. Lin X, Xu Y, Pan X, et al. Global, regional, and national burden and trend of diabetes in 195 countries and territories: an analysis from 1990 to 2025. Scientific Reports. 2020;10(1):14790.

2. Cho NH, Shaw J, Karuranga S, et al. IDF Diabetes Atlas: Global estimates of diabetes prevalence for 2017 and projections for 2045. Diabetes research and clinical practice. 2018;138:271–281.

3. Li J, Tong Y, Zhang Y, et al. Effects on All-cause Mortality and Cardiovascular Outcomes in Patients With Type 2 Diabetes by Comparing Insulin With Oral Hypoglycemic Agent Therapy: A Meta-analysis of Randomized Controlled Trials. Clinical Therapeutics. 2016;38(2):372–386.e376.

4. Liu J, Ren Z-H, Qiang H, et al. Trends in the incidence of diabetes mellitus: results from the Global Burden of Disease Study 2017 and implications for diabetes mellitus prevention. BMC Public Health. 2020;20(1):1415.

5. Williams R, Karuranga S, Malanda B, et al. Global and regional estimates and projections of diabetes-related health expenditure: Results from the International Diabetes Federation Diabetes Atlas, 9th edition. Diabetes Research and Clinical Practice. 2020;162.

6. van den Arend IJM, Stolk RP, Krans HMJ, Grobbee DE, Schrijvers AJP. Management of type 2 diabetes: a challenge for patient and physician. Patient Education and Counseling. 2000;40(2):187–194.

7. Cetin M, Sahin S. Microparticulate and nanoparticulate drug delivery systems for metformin hydrochloride. Drug Delivery. 2016;23(8):2796–2805.

8. McCoy RG, Dykhoff HJ, Sangaralingham L, et al. Adoption of new glucose-lowering medications in the US—the case of SGLT2 inhibitors: nationwide cohort study. 2019;21(12):702–712.

9. Nauck MA, Wefers J, Meier JJJTLD, Endocrinology. Treatment of type 2 diabetes: challenges, hopes, and anticipated successes. 2021;9(8):525–544.

10. Crush K. Carnosine and related substances in animal tissues. Comparative biochemistry and physiology. 1970;34(1):3–30.

11. Kohen R, Yamamoto Y, Cundy KC, Ames BN. Antioxidant activity of carnosine, homocarnosine, and anserine present in muscle and brain. Proceedings of the National Academy of Sciences. 1988;85(9):3175–3179.

12. Peters V, Klessens CQF, Baelde HJ, et al. Intrinsic carnosine metabolism in the human kidney. Amino Acids. 2015;47(12):2541–2550.

13. Matthews JJ, Artioli GG, Turner MD, Sale C. The physiological roles of carnosine and β-alanine in exercising human skeletal muscle. Medicine & Science in Sports & Exercise. 2019;51(10):2098–2108.

14. Baye E, Ukropcova B, Ukropec J, Hipkiss A, Aldini G, de Courten B. Physiological and therapeutic effects of carnosine on cardiometabolic risk and disease. Amino Acids. 2016;48(5):1131–1149.

15. Feehan J, Hariharan R, Buckenham T, et al. Carnosine as a potential therapeutic for the management of peripheral vascular disease. 2022.

16. de Courten B, Jakubova M, de Courten MP, et al. Effects of carnosine supplementation on glucose metabolism: Pilot clinical trial. Obesity. 2016;24(5):1027–1034.

17. Houjeghani S, Kheirouri S, Faraji E, Jafarabadi MA. l-Carnosine supplementation attenuated fasting glucose, triglycerides, advanced glycation end products, and tumor necrosis factor–α levels in patients with type 2 diabetes: a double-blind placebo-controlled randomized clinical trial. Nutrition Research. 2018;49:96–106.

18. Elbarbary NS, Ismail EAR, El-Naggar AR, Hamouda MH, El-Hamamsy M. The effect of 12 weeks carnosine supplementation on renal functional integrity and oxidative stress in pediatric patients with diabetic nephropathy: a randomized placebo-controlled trial. Pediatr Diabetes. 2018;19(3):470–477.

19. Nealon R, Sukala W, Coutts R, Zhou S. The effect of 28 days of beta-alanine supplementation on exercise capacity and insulin sensitivity in individuals with type 2 diabetes mellitus: a randomised, double-blind and placebo-controlled pilot trial. J Nutr Sci Res. 2016;1(3):1–7.

20. Baye E, Ukropec J, de Courten MPJ, et al. Effect of carnosine supplementation on the plasma lipidome in overweight and obese adults: a pilot randomised controlled trial. Scientific Reports. 2017;7(1):17458.

21. Erion DM, Park HJ, Lee HY. The role of lipids in the pathogenesis and treatment of type 2 diabetes and associated co-morbidities. BMB Rep. 2016;49(3):139–148.

22. Parhofer KG. Interaction between Glucose and Lipid Metabolism: More than Diabetic Dyslipidemia. Diabetes Metab J. 2015;39(5):353–362.

23. Ashtary-Larky D, Bagheri R, Ghanavati M, et al. Effects of beta-alanine supplementation on body composition: a GRADE-assessed systematic review and meta-analysis. Journal of the International Society of Sports Nutrition. 2022;19(1):196–218.

24. Chan A-W, Tetzlaff JM, Gøtzsche PC, et al. SPIRIT 2013 explanation and elaboration: guidance for protocols of clinical trials. Bmj. 2013;346.

25. Schulz KF, Altman DG, Moher D. CONSORT 2010 Statement: Updated Guidelines for Reporting Parallel Group Randomized Trials. Annals of Internal Medicine. 2010;152(11):726–732.

26. World Medical Association Declaration of Helsinki: ethical principles for medical research involving human subjects. Jama. 2013;310(20):2191–2194.

27. Menon K, Cameron JD, de Courten M, de Courten B. Use of carnosine in the prevention of cardiometabolic risk factors in overweight and obese individuals: study protocol for a randomised, double-blind placebo-controlled trial. BMJ open. 2021;11(5):e043680–e043680.

28. Baye E, Menon K, de Courten MPJ, Earnest A, Cameron J, de Courten B. Does supplementation with carnosine improve cardiometabolic health and cognitive function in patients with pre-diabetes and type 2 diabetes? study protocol for a randomised, doubleblind, placebo-controlled trial. BMJ Open. 2017;7(9):e017691.

29. Matthews DR, Hosker JP, Rudenski AS, Naylor BA, Treacher DF, Turner RC. Homeostasis model assessment: insulin resistance and β-cell function from fasting plasma glucose and insulin concentrations in man. Diabetologia. 1985;28(7):412–419.

30. Menon K, de Courten B, Magliano DJ, Ademi Z, Liew D, Zomer E. The Cost-Effectiveness of Supplemental Carnosine in Type 2 Diabetes. Nutrients. 2022;14(1):215.

31. Riedel AA, Heien H, Wogen J, Plauschinat CA. Loss of Glycemic Control in Patients with Type 2 Diabetes Mellitus Who Were Receiving Initial Metformin, Sulfonylurea, or Thiazolidinedione Monotherapy. Pharmacotherapy: The Journal of Human Pharmacology and Drug Therapy. 2007;27(8):1102–1110.

32. Perreault L, Pan Q, Mather KJ, Watson KE, Hamman RF, Kahn SE. Effect of regression from prediabetes to normal glucose regulation on long-term reduction in diabetes risk: results from the Diabetes Prevention Program Outcomes Study. The Lancet. 2012;379(9833):2243–2251.

33. Menon K, Marquina C, Liew D, Mousa A, de Courten B. Histidine-containing dipeptides reduce central obesity and improve glycaemic outcomes: A systematic review and metaanalysis of randomized controlled trials. Obesity Reviews. 2020;21(3):e12975.

34. Matthews JJ, Dolan E, Swinton PA, et al. Effect of Carnosine or β-Alanine Supplementation on Markers of Glycemic Control and Insulin Resistance in Humans and Animals: A Systematic Review and Meta-analysis. Advances in Nutrition. 2021;12(6):2216–2231.

35. Elbarbary NS, Ismail EAR, El-Naggar AR, Hamouda MH, El-Hamamsy M. The effect of 12 weeks carnosine supplementation on renal functional integrity and oxidative stress in pediatric patients with diabetic nephropathy: a randomized placebo-controlled trial. Pediatric Diabetes. 2018;19(3):470–477.

36. Liu Y, Cotillard A, Vatier C, et al. Correction: A Dietary Supplement Containing Cinnamon, Chromium and Carnosine Decreases Fasting Plasma Glucose and Increases Lean Mass in Overweight or Obese Pre-Diabetic Subjects: A Randomized, Placebo-Controlled Trial. Plos one. 2015;10(12):e0145315.

37. Everaert I, Taes Y, Heer ED, et al. Low plasma carnosinase activity promotes carnosinemia after carnosine ingestion in humans. American Journal of Physiology-Renal Physiology. 2012;302(12):F1537–F1544.

38. Kresta JY, Oliver JM, Jagim AR, et al. Effects of 28 days of beta-alanine and creatine supplementation on muscle carnosine, body composition and exercise performance in recreationally active females. Journal of the International Society of Sports Nutrition. 2014;11(1):55.

39. Kendrick IP, Harris RC, Kim HJ, et al. The effects of 10 weeks of resistance training combined with β-alanine supplementation on whole body strength, force production, muscular endurance and body composition. Amino Acids. 2008;34(4):547–554.

40. Regazzoni L, de Courten B, Garzon D, et al. A carnosine intervention study in overweight human volunteers: bioavailability and reactive carbonyl species sequestering effect. Scientific reports. 2016;6:27224.

41. Meerwaldt R, Links T, Zeebregts C, Tio R, Hillebrands J-L, Smit A. The clinical relevance of assessing advanced glycation endproducts accumulation in diabetes. Cardiovascular Diabetology. 2008;7(1):29.

42. de Courten B, de Courten MP, Soldatos G, et al. Diet low in advanced glycation end products increases insulin sensitivity in healthy overweight individuals: a double-blind, randomized, crossover trial. Am J Clin Nutr. 2016;103(6):1426–1433.

43. Reddy VP, Garrett MR, Perry G, Smith MA. Carnosine: A Versatile Antioxidant and Antiglycating Agent. Science of Aging Knowledge Environment. 2005;2005(18):pe12–pe12.

44. Hipkiss AR, Chana H. Carnosine Protects Proteins against Methylglyoxal-Mediated Modifications. Biochemical and Biophysical Research Communications. 1998;248(1):28–32.

45. Maillard LC. Action des acides amines sur les sucres: formation des melanoidines par voie methodique. CR Acad Sci. 1912;154:66–68.

46. Hobart LJ, Seibel I, Yeargans GS, Seidler NW. Anti-crosslinking properties of carnosine: Significance of histidine. Life Sciences. 2004;75(11):1379–1389.

47. Brownlee M, Pongor S, Cerami A. Covalent attachment of soluble proteins by nonenzymatically glycosylated collagen. Role in the in situ formation of immune complexes. The Journal of experimental medicine. 1983;158(5):1739–1744.

48. Baye E, Ukropec J, De Courten MP, et al. Effect of carnosine supplementation on the plasma lipidome in overweight and obese adults: a pilot randomised controlled trial. 2017;7(1):1–7.

49. Menon K, Marquina C, Hoj P, Liew D, Mousa A, de Courten B. Carnosine and histidine-containing dipeptides improve dyslipidemia: a systematic review and meta-analysis of randomized controlled trials. Nutrition Reviews. 2020;78(11):939–951.

